# Preclinical Efficacy in Investigator’s Brochures: Stakeholders’ views on measures to improve Completeness and Robustness

**DOI:** 10.1101/2022.02.07.22270434

**Authors:** Martin Haslberger, Susanne Gabriele Schorr, Daniel Strech, Tamarinde Haven

## Abstract

Research ethics committees (RECs) and regulatory agencies assess whether the benefits of a proposed early-stage clinical trial outweigh the risks based on preclinical studies reported in investigator’s brochures (IBs). Recent studies have indicated that preclinical evidence presented in IBs is reported in a way that does not enable proper risk-benefit assessment. We interviewed different stakeholders (regulators, REC members, industry representatives, preclinical and clinical researchers, ethicists, and metaresearchers) about their views on measures to increase the completeness and robustness of preclinical evidence reporting in IBs.

This study was preregistered (https://osf.io/nvzwy/). We used purposive sampling and invited stakeholders to participate in an online semistructured interview between March and June 2021. The themes were derived using inductive content analysis. We used a strengths, weaknesses, opportunities, and threats (SWOT) matrix to categorize our findings.

Twenty-seven international stakeholders participated. The interviewees pointed to several strengths and opportunities to improve completeness and robustness, mainly more transparent and systematic justifications of the inclusion of studies. However, weaknesses and threats were mentioned that could undermine efforts to enable more thorough assessment: The interviewees stressed that current review practices are sufficient to ensure the safe conduct of first-in-human trials. They feared that changes to the IB structure or review process could overburden stakeholders and slow drug development.

In principle, having more robust decision-making processes in place aligns with the interests of all stakeholders and with many current initiatives to increase the translatability of preclinical research and limit uninformative or ill-justified trials early in the development process. Further research should investigate measures that could be implemented to benefit all stakeholders.

## INTRODUCTION

To ensure that clinical trials are ethical, foundational ethical documents require regulatory agencies and research ethics committees (RECs) to perform a risk-benefit assessment (RBA) based on supporting evidence (1,2). In practice, this RBA is based on studies reported in investigator’s brochures (IBs). IBs present the collected preclinical and clinical evidence relevant to the study of an investigational product in human subjects (3). Preclinical studies are an important form of decision support for early-phase clinical trial approvals, making the IB a critical document in translation.

Recent studies have indicated that preclinical and clinical evidence supporting the study rationale in IBs is reported in a way that does not enable proper RBA – one study found that the majority of IBs (82%) described only positive findings for preclinical efficacy, raising concerns about potential reporting biases (4).

Items to assess the risk of bias in the included studies were routinely missing, and little insight was provided into how the presented evidence was compiled, which raised further concerns about the reporting and design biases of the included studies (4–6). The authors argued that if the current guidance regarding evidence reporting in IBs is too vague, there is little opportunity for RECs and other stakeholders to play their critical gatekeeping role in the translation process. This argument seems to be supported by academic groups investigating the evidence base underlying early-stage trials (7,8). Issues raised in these publications include the improper selection of studies and alleged misrepresentation of animal data. These issues could result in human subjects being exposed to unnecessary risks associated with experimental compounds when the supporting studies did not warrant translation, which would contradict fundamental ethical principles and regulatory guidance (2,3).

Concerns about the rigor and reporting of preclinical studies have been widely published (9–13). They have been echoed by medical ethicists arguing that more emphasis should be placed on the preclinical promise of efficacy before conducting first trials in human subjects. A greater focus on the validity and complete reporting of preclinical efficacy studies used to make regulatory decisions to translate could be a possible lever for reducing possible blind spots in ethical and scientific review and better protecting research participants (14).

We aim here to compile stakeholders’ (regulators, REC members, industry representatives, preclinical and clinical researchers, ethicists, and metaresearchers) views on two key concerns: [1] transparent criteria for evidence selection and synthesis to ensure the completeness of all preclinical evidence for efficacy presented in IBs and [2] the relevance of improving the reporting of items to assess the robustness of studies. We unpack these questions by determining the strengths, weaknesses, opportunities, and threats (SWOT) of those concerns and proposed measures to address them. Finally, we reflect on whether or how our interviewees believed these suggestions could be implemented.

## METHODS

### Ethical approval

The Charité University Medical Center committee reviewed and approved our study protocol under application number EA4/026/21. The participants provided written informed consent.

### Participants

We used purposive sampling (15) to gain perspectives from stakeholders involved in compiling or reviewing IBs. In addition, we solicited views from metaresearchers and ethicists with a focus on translational research.

We recruited participants in three separate ways: a) through our own networks, b) through internet searches based on the participants’ functions (i.e., cold emailing) and c) through snowballing (recommendations by interviewees or pilot participants). We aimed to recruit at least 5 interviewees from each major stakeholder group to obtain a sufficiently broad overview of stakeholder perspectives.

### Procedure

This study was preregistered on the Open Science Framework (https://osf.io/nvzwy/). We invited the participants via e-mail and, if we did not receive a response, reminded them within 10 days. We attached the information letter and informed consent form to the e-mail and included a link to the study protocol. Interviewees who requested access to the interview questions prior to the interview received a shortened version of the topic guide. The interviews were conducted via video call between March and June 2021 and took between 25 and 50 minutes. Two team members attended each interview (TH and MH), one as an interviewer and one for note-taking and technical support. DS and TH had previously designed and carried out various qualitative studies and trained MH in qualitative research methodology. The interviewers met after each interview for peer debriefing. The interviews were transcribed ad verbatim by a transcription company and proofread by TH or MH prior to analysis. Further information on the procedure can be found in our protocol (https://osf.io/4msje/).

### Interview structure

We performed initial PubMed and Google searches to inform an internal discussion on potential topics for the interviews and associated pros and cons. The discussion resulted in a topic guide (https://osf.io/mjbv2/) to outline the interview structure. We tested the topic guide with colleagues (n=4) using cognitive interviewing, which resulted in minor changes for comprehensibility. After a brief introduction of the topic, we started each interview with a question about the role that IBs played in the interviewees’ work. Then, we enquired about the interviewees’ views on the relevance of improving the reporting of robustness items and the completeness of preclinical evidence in IBs and asked for suggestions to improve the structure or assessment of IBs. When relevant, we discussed measures based on previous interviews or the literature search (examples are included in the topic guide). Finally, we offered the interviewees a written summary of the interview with the option to comment or correct (member check). If they agreed, we sent the summary via e-mail and integrated their corrections into our analysis.

### Analysis

We used MAXQDA (Release 20.4.0) for coding and data analysis. We analyzed the interviews with a combination of deductive and inductive content analysis (16). The main structure of the analysis was a SWOT matrix (deductive), but subthemes within the SWOT matrix (see Table 1) were identified inductively. Our analysis consisted of four steps. First, two team members (MH and TH) read and analyzed the same 5 transcripts independently and used in vivo coding and clustering to identify subthemes. Second, the two team members (MH and TH) met and discussed their codes, resolving any disagreements through discussion and refining code descriptions. Third, the code tree based on the discussion was discussed among all the authors, which resulted in minor modifications. Fourth, MH and TH read and coded the remaining transcripts with this coding tree until thematic saturation was reached (17), meaning that some small modifications were made but no novel codes were identified. The final code tree is a SWOT matrix of measures to improve the completeness and robustness of reporting of supporting evidence for efficacy in IBs. We used the COREQ reporting guideline (18) to structure our findings.

**Table 1:** SWOT matrix definitions for increased attention to preclinical efficacy in early clinical research

|  |  |
| --- | --- |
| Strengths | Arguments in favor of increased attention to robustness and completeness of preclinical evidence for efficacy |
| Weaknesses | Arguments against increased attention to preclinical efficacy and potential negative effects |
| Opportunities | Potential long-term effects of changes in review practices or the regulatory ecosystem that would facilitate a more streamlined and thorough review of robustness and completeness |
| Threats | Circumstances that could pose a barrier to possible changes in the regulatory ecosystem or hinder the implementation of more stringent guidance or requirements |

## RESULTS

### Interviews

We contacted 64 people with a success rate of 42%, resulting in a total of 25 interviews with 27 participants (one interview was with three interviewees) between March and June 2021 (see Figure 1). In total, 4 participants (15%) were recruited through our own network, 15 (56%) were recruited through snowballing, and 8 (30%) were invited based on their relevant experience but had no connection to the research team. Of our interviewees, 19 (70%) indicated that they regularly worked with IBs, and most of the others – ethicists, policy makers, or methodological researchers – had previous experience with IBs. Nineteen interviewees (70%) requested member checks, and we integrated their corrections prior to analysis.

**Figure 1:**
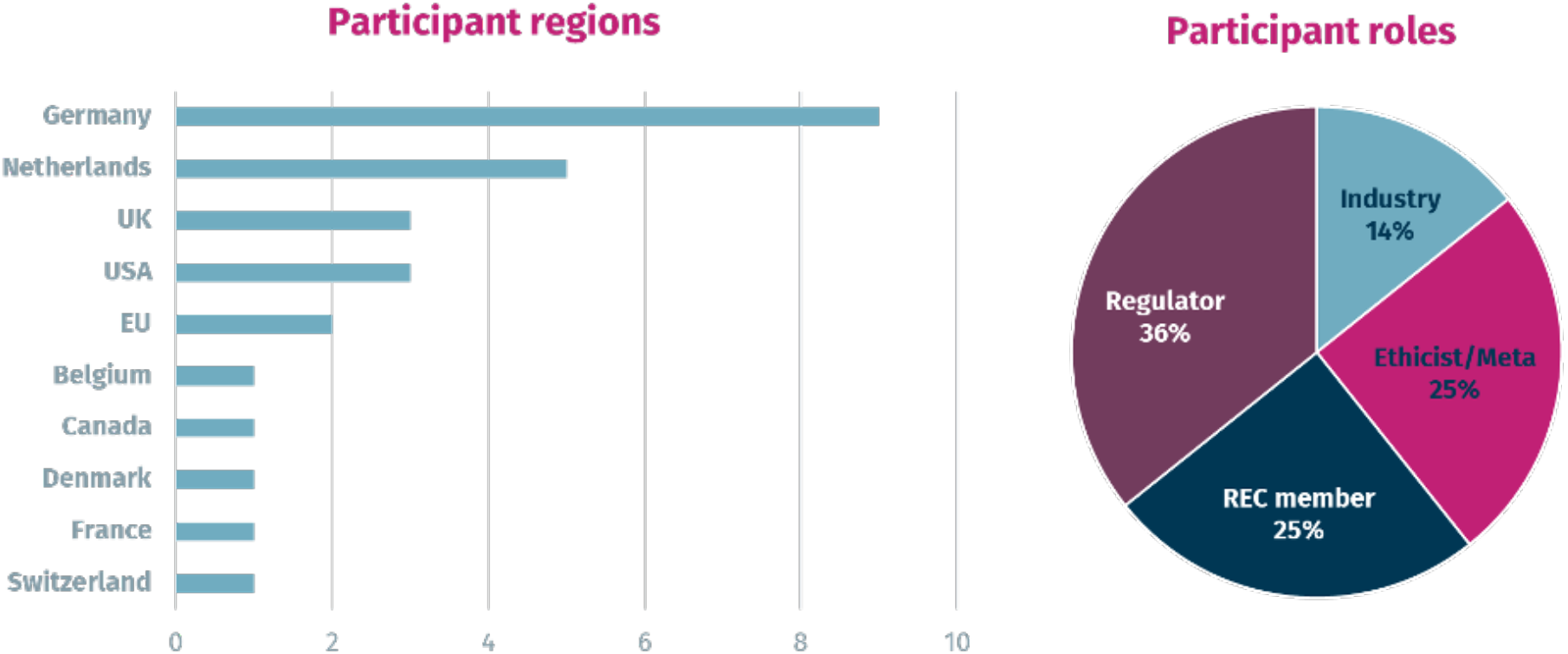
Participant characteristics. A total of 27 people participated. A more detailed breakdown is available in supplementary Table S1.

**Figure 2:**
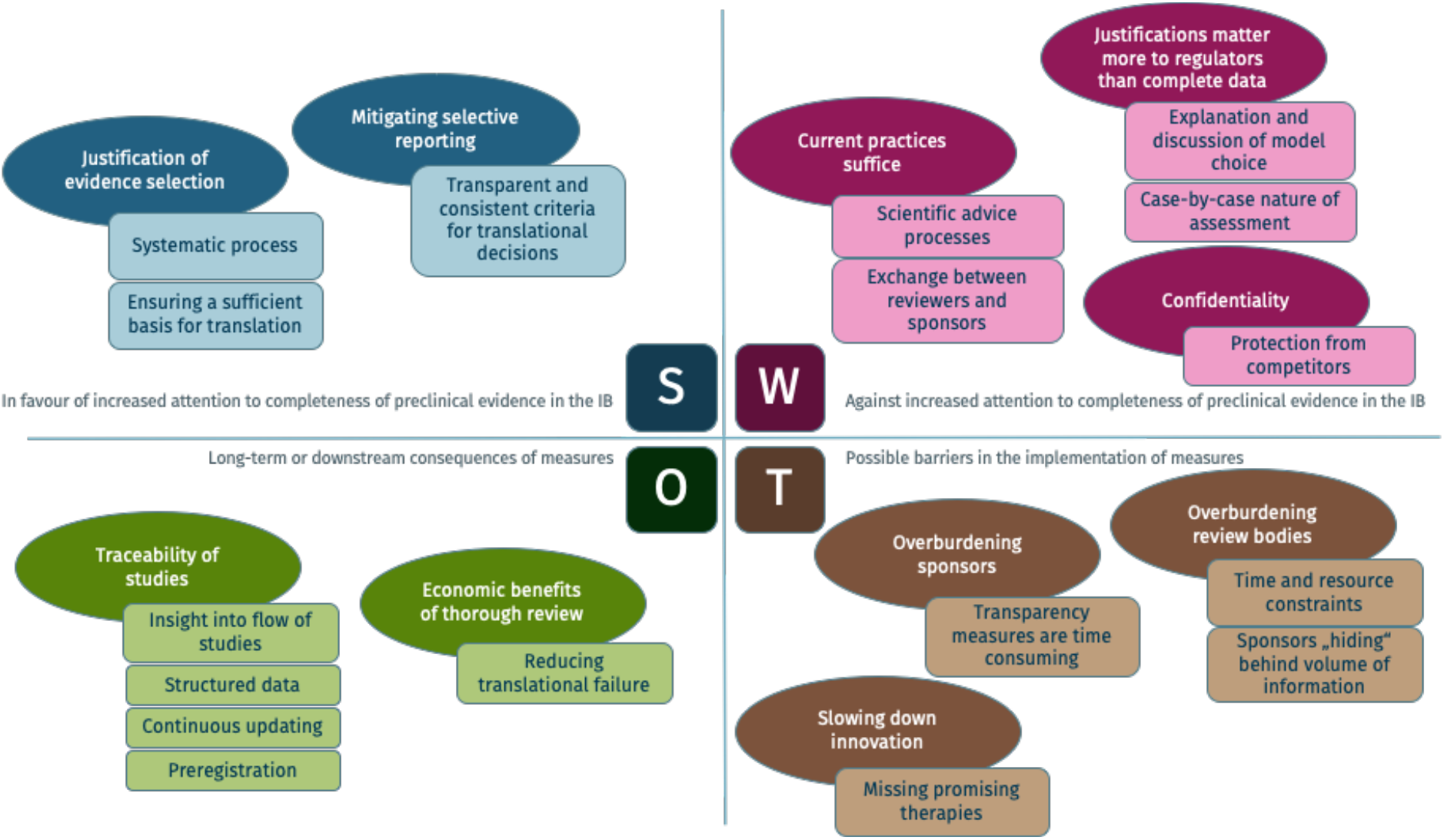
SWOT matrix visualization of increased attention to complete reporting of all preclinical efficacy studies in early clinical research. Darker shaded ovals represent overarching themes, and lighter shaded boxes represent subthemes.

**Figure 3:**
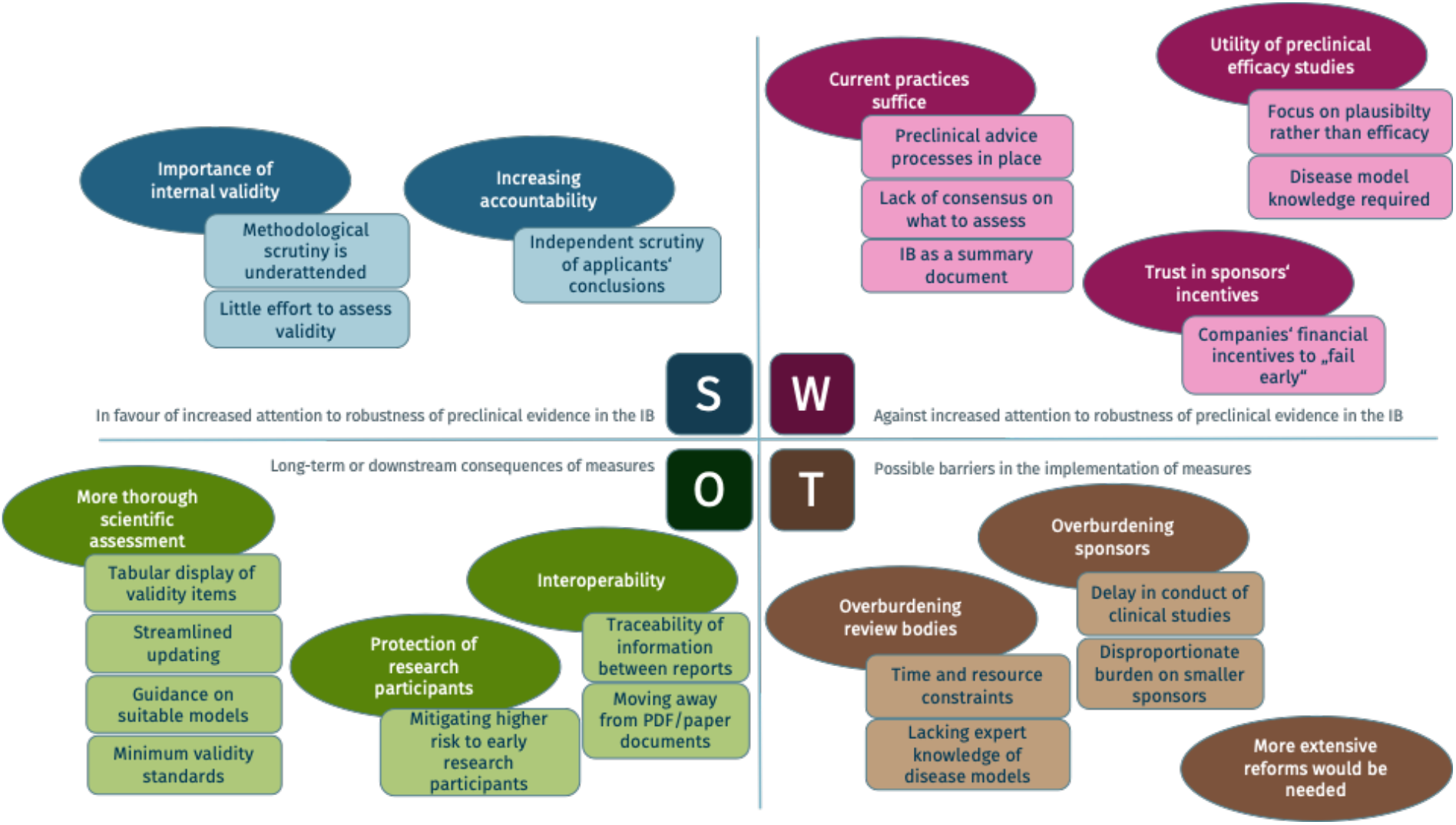
SWOT matrix visualization of increased attention to the robustness of preclinical efficacy studies in early clinical research. Darker shaded ovals represent overarching themes, and lighter shaded boxes represent subthemes.

### SWOT analysis

We used a SWOT matrix to structure our codes. The two main interview topics – completeness and robustness of data – were addressed separately in the topic guide. The definitions used for the SWOT items are shown in Table 1.

Quotations exemplifying the themes presented in this section are displayed in supplementary Tables S2 (Completeness) and S3 (Robustness).

### Completeness

### POTENTIAL STRENGTHS

#### Justification of evidence selection (Table S2, Q. 1–3)

Members of both RECs and regulatory authorities reported relying on the IB instead of their own literature searches to assess the supporting evidence of trial applications. The interviewees indicated that they would appreciate increased attention to the completeness of preclinical efficacy data for multiple reasons:

The interviewees mentioned that presenting the complete underlying data is a non-negotiable requirement for conducting a proposed clinical trial and is implicit in the available ethical and regulatory guidance. Some interviewees, particularly pharmacologists and REC members, mentioned that they were not aware of or had never considered the possibility of publication or design biases in the IB. Others pointed out that sponsors construct the IB with the end goal of obtaining approval and that studies that do not support that goal might be left out.

Interviewees across groups pointed out that in principle, all stakeholders involved in the review should have access to the complete existing evidence, including that in the public domain and studies conducted in house by the sponsor.

Different interviewees pointed out that all available relevant data should be presented and exclusions should be justified in a transparent and concise manner. Metaresearcher and ethicist participants mentioned that systematic reviews would ideally be conducted by an independent entity. Some regulators mentioned that they occasionally perform literature searches themselves to check whether additional data can be found in the public domain.

#### Mitigating selective reporting(Table S2, Q. 4–7)

As preclinical studies are often proprietary and not publicly available, sponsors could in theory choose to selectively report studies without verification by regulatory gatekeepers. Similarly, regulators and RECs do not have the resources to routinely check the public domain. Some interviewees mentioned that such selective reporting would conflict with sponsors’ goals, as it is very expensive to drag out the failure of an investigational product with a lack of evidentiary support to the late clinical phases. However, some were concerned that individual motives can conflict with this “fail early” theme: individuals who are “believers” in their compounds or companies that have a financial interest in exaggerating the products in their pipeline (e.g., before a company sale or initial public offering) might be more susceptible to selectively reporting positive studies and distorting the picture that is presented to regulators, RECs, and the public. Similarly, academic research funded by industry could be at higher risk of bias. Interviewees also mentioned that decision making in pharmaceutical companies is decoupled from evidence presented in the IB, which might be seen as a tool for passing a regulatory hurdle but is not a true representation of the internal decision-making process.

### POTENTIAL WEAKNESSES

#### Justifications matter more to regulators than complete data(Table S2, Q. 8–9)

Different regulators pointed out that they value receiving the right justifications for why certain studies were performed and explanations of their meaning and limitations rather than having a complete picture of all conducted studies. Some mentioned that their point of view during assessment is “Are the data enough to justify this trial?” rather than “Are these all data that are available?”

Interviewees across groups cautioned that subjecting the IB to many structural provisions would not be feasible, as such restrictions might not accommodate the large variety of proposals. Some indications might not allow for primary pharmacology data to be obtained due to a lack of fitting disease models, or the data for different proposals could be too heterogeneous to be organized in the same way or could be field specific. Some interviewees representing regulatory agencies noted that they make decisions on a case-by-case basis and prefer the relevant data to be presented concisely, but not necessarily in a more structured format.

#### Confidentiality/competitive aspects (Table S2, Q. 10–13)

Interviewees noted that the efficacy studies presented in IBs can be confidential, which could undermine the usefulness of some of the proposed measures to improve completeness.

Pharmaceutical companies might object to providing insight into their proprietary data because they wish to protect those data from competitors.

#### Current regulatory advice and assessment practices suffice

Some interviewees considered additional measures superfluous in the face of current practices. Various REC members reported that if serious issues with a protocol or IB arose, they would ask sponsors for more data. However, this process was described as very time intensive, and RECs and regulatory agencies lack the resources to do it routinely. Regulators pointed out that they offer scientific advice to sponsors on the studies needed for regulatory purposes from an earlier development stage onward; however, they rarely ask for additional efficacy studies to be performed.

### POTENTIAL OPPORTUNITIES

#### Traceability of studies (Table S2, Q. 13–14)

Interviewees highlighted that with greater emphasis on how evidence was compiled, the process of justifying a trial becomes more traceable. In addition, some noted that Ibs that are mainly text based can have limited expressive power and that including tables showing all studies or a standardized set of key plots could make the assessment more efficient. Other suggestions included visualizations known from systematic reviews, such as forest plots and graphics to provide insight into the flow of reasoning between studies.

Some remarked that moving away from PDF documents and toward structured data files could enable more frequent updating of Ibs and be a clearer way to package new evidence as new studies are conducted during the drug development process.

A few interviewees commented that to mitigate the possibility of selective reporting, the best solution would be for reviewers to require that all presented preclinical data be preregistered. This would enable actual checks of whether studies were excluded. However, such registries would have to encompass all preclinical research globally to include the various data sources that may be cited in an IB, including all academic and industry research.

#### Economic benefits of thorough review (Table S2, Q. 15–16)

Interviewees pointed to the estimates of research spending wasted on uninformative trials and irreproducible research to highlight the advantage of a thorough scientific review, which could reduce translational failures.

### POTENTIAL THREATS

#### Overburdening review bodies (Table S2, Q. 17–18)

Imposing additional responsibilities on regulators or RECs to perform checks of the completeness of studies or adding sections describing the evidence synthesis without providing more resources could overburden them due to time constraints. Relatedly, regulators cautioned that they might be overwhelmed by the inclusion of too many studies when only those most relevant to the proposal are needed. One regulator mentioned that should they, in contrast to the current practice, see a more balanced picture of the underlying evidence that includes a larger number of negative studies, this could even be considered a red flag and put a stop to research that might otherwise have been approved. Similarly, another noted that an overly broad inclusion of studies would currently raise suspicions, as it might be interpreted as the sponsor “hiding” information in the mass of other studies.

#### Overburdening sponsors

Some interviewees highlighted that additional efforts to increase the transparency of the evidence selection might be more time consuming and therefore burden sponsors, particularly small companies and academic sponsors.

#### Slowing innovation (Table S2, Q. 19)

Interviewees warned that having overly strict criteria for which types of studies to include to show efficacy could be a potential risk. They reasoned that this restriction might lead to overlooking promising therapies for which no disease model yet exists or for which only biomarkers of effect with limited clinical value have been found.

### Robustness

### POTENTIAL STRENGTHS

#### Importance of internal validity of experiments (Table S3, Q. 1–4)

The interviewees emphasized that the robustness of preclinical studies is an important parameter in a clinical trial application (CTA). Some pointed to ongoing discussions about the value of preclinical research; more focus on the internal validity and greater methodological scrutiny of preclinical studies could be an area of improvement that is currently neglected. Interviewees in reviewer positions noted that the robustness of pharmacodynamics studies is often weak. Historically, regulators have not assigned much weight to robustness and trusted that the data were robust, as they reasoned that otherwise, the sponsors would not proceed with clinical development.

Interviewees noted that items to assess robustness threats, e.g., the “Landis-4” criteria (sample size estimation, randomization, blinded outcome assessment, and handling of data), can be presented concisely, e.g., by tabulating all studies with their corresponding values. This would require little extra effort from sponsors and could allow a quick grading of the evidence by reviewers.

#### Increasing accountability (Table S3, Q. 5–6)

The industry representatives assumed that regulators assess the concordance between the IB contents and the underlying preclinical study reports. The regulators reported that this assessment as well as an in-depth review of the validity of the study reports is possible in theory. However, doing so routinely would be untenable for them due to tight deadlines, and they would do so only if they spotted red flags. Because of the tight deadlines, the regulators said that they need to rely on the conclusions drawn by the applicants.

### POTENTIAL WEAKNESSES

#### Current assessment and advice processes suffice (Table S3, Q. 7–10)

Regulators pointed out that sponsors are often in contact with them before submitting a CTA and discuss concrete requirements within scientific advice processes. Additionally, regulators mentioned having access to underlying study reports and that REC members are granted access to reports upon request. Some REC members and regulators reported initiating exchanges with sponsors to obtain more insight into the sponsors’ evaluations of the presented evidence. However, to what extent this process captures advice on the robustness of studies was not specified.

Interviewees noted that, in contrast to GLP for safety/toxicology studies (although it also does not cover internal validity), there is only limited guidance on the conduct of efficacy studies. While the most recent EMA guidance on first-in-human trials pointed to increased discussion of the models used in efficacy studies, there are no specific requirements for their robustness. Additionally, evidence presented for efficacy might consist only of literature references with no information on validity threats.

Various interviewees mentioned that the IB as a summary document is not suitable for the detailed reporting of study characteristics and that such reporting requirements would be difficult to capture in the format or would inflate the IB.

#### Trust in sponsors’ incentives (Table S3, Q. 11)

Interviewees from all groups noted that sponsors have strong financial incentives to terminate unpromising development projects early. Regulators and REC members assumed that pharmaceutical companies have internal robustness checks in place and are aware of the quality and robustness of different disease models.

#### Utility of preclinical efficacy studies (Table S3, Q. 12–14)

Several interviewees objected more generally to the notion that efficacy is a very important parameter in preclinical development. They raised various interrelated points regarding this issue. First, preclinical studies are used to assess predictable effects in humans. Most animal studies are used to obtain a proof of concept for a mechanism of action, but they are generally considered to have little predictive value for clinical efficacy. Second, many of the interviewees generally focused their assessment of preclinical data on the plausibility of pathophysiological concepts or proposed mechanisms of action and did not agree that internal validity should play an emphasized role in the assessment. Third, some of the participants pointed to the complete lack of suitable animal models for certain indications and argued that it would be preferable not to conduct experiments in those cases.

### POTENTIAL OPPORTUNITIES

#### Explicit deliverables could enable a more thorough scientific assessment (Table S3, Q. 15–17)

Some interviewees noted that an explicit set of deliverables to include in an IB and against which to assess the robustness of preclinical evidence would be welcomed. In particular, regulators and EC pharmacologists argued that standardized tabular overviews of the conducted studies would be a very direct improvement to obtain more consistency in how data are presented. These tables could include validity items (e.g., “Landis-4”). They could allow for more efficient assessment by oversight bodies and save time by better integrating the studies with existing software tools used by pharmacologists for dose finding. They could also save time when updating the IB, as less text would have to be produced, and new studies over the course of development, including items used to assess their robustness, could be added to the list. Other participants called for a certification of nonregulated research, similar to the GLP mechanics. Some interviewees also called for clearer guidance on which disease models are suitable to present as preclinical efficacy studies.

#### Interoperability (Table S3, Q. 18–21)

The interviewees saw opportunities in ensuring that the preclinical information would be more traceable between the original publication (study report, publication, etc.) and the IB. This could entail moving away from the IB as a paper document and toward a more interactive assessment. A potential way to achieve this would be through a software solution where the reviewers themselves could create summaries of the supporting evidence as needed. Some proposed building such software on top of the structured electronic submission of original data (e.g., CDISC’s Standard for Exchange of Nonclinical Data (SEND) format). This could allow for an assessment of trial application documents at different depths, based on the level of scrutiny needed, down to the level of individual study reports.

#### Protection of research participants (Table S3, Q. 22–24)

Regulators remarked that much less time and attention are given to applications for clinical trials compared to marketing authorizations, which they found odd considering the higher level of uncertainty and risk associated with (mostly healthy volunteer) trial participants.

Some interviewees, mainly ethicists and methodologists, proposed that more extensive changes in the research ecosystem would be necessary to assure the trustworthiness of the preclinical evidence base. One suggestion was to set stringent minimum validity requirements for studies to be included in an IB. The otherwise negatively framed notion of such measures possibly bringing fewer innovative therapies into the clinic was framed positively here: the interviewees alluded to an overextension of the freedom of research that is granted in the preclinical realm to early human research and thought that more conservative practices could fit with the “fail early” paradigm in drug development.

### POTENTIAL THREATS

#### Overburdening review bodies (Table S3, Q. 25)

Interviewees were most concerned about the time and resource constraints for all stakeholders involved in the review of IBs. RECs must rely on recruiting qualified pharmacologists, who were described as increasingly hard to find. The overall scope of the ethical review in an EC is centered around the protocol, not the IB – REC members said that they would be overwhelmed, as they do not have the resources to review the IB thoroughly. Among the REC members, only the pharmacologists/pharmacists reported reviewing the IB of an application. Some interviewees criticized this review as an excessive amount of responsibility for a single committee member. Additionally, clinical trial offices in national regulatory authorities and RECs in most countries assess applications from a range of indications. This makes it unlikely that there is an expert available for every indication who could assess the validity of the models presented in the applications in detail.

#### Overburdening sponsors (Table S3, Q. 26–27)

Interviewees pointed out that overburdening sponsors with more specific robustness requirements would hinder or delay the conduct of exploratory trials. This could slow innovation and make research more expensive. Some also mentioned that a more formal scientific assessment from RECs in addition to the assessment by regulatory agencies would constitute an unnecessary double regulatory hurdle for sponsors that could slow the progress of clinical research. They pointed out that stronger regulation would disproportionately harm smaller companies or academic sponsors, as they often do not have the same experience or resources as larger companies.

#### More extensive reforms would be needed

Finally, various interviewees stressed that improving reporting or document structure alone might result in little real improvement. They raised the point that improvement should reside not only in how the evidence is reported but also in how preclinical research is conducted and the resulting evidence is reviewed. Without a shared endorsement of good research and reporting practices between stakeholders, matching incentive structures, and increased requirements, increased guidance might only increase the burden for all stakeholders.

## DISCUSSION

We gathered stakeholders’ views on measures to improve the completeness of evidence and the reporting of robustness data for preclinical efficacy in IBs. Overall, the interviewees appreciated greater attention to completeness and robustness. They noted that a more thorough justification for the selection of evidence could be helpful, especially in light of concerns over selective reporting. Similarly, the stakeholders recognized the importance of robustness and internal validity. However, they pointed to a lack of guidance for assessing the robustness of preclinical evidence for efficacy. Various stakeholders were concerned that placing greater emphasis on completeness and robustness would overburden the system. Some explicitly questioned the necessity of changing the reporting of preclinical evidence in IBs, as the system seems to have worked reasonably well in the past. In addition, interviewees underscored that stakeholders depend on each other to assure that the evidence is as complete and robust as possible.

### Contextualization

Stakeholders are increasingly recognizing the need for greater attention to the robustness of preclinical data (9–13). Measures commented on or suggested by stakeholders to increase robustness included the reporting of internal validity items for key studies in tabulated form, minimum validity standards and quality certifications. Governmental funders of basic research, such as the National Institutes of Health in the United States, now also emphasize the importance of internal validity in preclinical research (19). Another example is industry-academia consortia developing quality standards for preclinical studies (20). In addition, European and North American regulators have included guidance on model relevance and animal species (21,22). The EMA first-in-human guideline explicitly mentions that studies influencing the design of CTs should be of high quality and reliability. Regulatory comments echo this statement (23).

While there have been proposals for the assessment of the robustness of preclinical evidence for RBA (14,24), a comparison of the therapeutic guidelines of the EMA and FDA found that a minority of the guidelines include recommendations or discussions of efficacy testing, even fewer specify suitable models, and none address issues of internal validity (25). In conclusion, guidance for assessing the robustness or internal validity of preclinical efficacy data exists in the academic literature, but official guidance from the EMA and FDA is still lacking. Such guidance could depart from a minimum set of validity criteria such as the “Landis-4” or ARRIVE “Essential 10” (11,13). Another option could be that studies included in the IB must be independently replicated (similar to some efforts in cancer biology (26,27)) or be explicitly designed as confirmatory studies (28).

Some interviewees were skeptical of increased attention to the robustness of preclinical efficacy data. They pointed out that the conduct of first-in-human studies has generally been safe, highlighting the focus on safety in early clinical development. Although conceptually separating safety and efficacy can be challenging, our focus was on efficacy and the determination of clinical promise based on preclinical efficacy data (14). The supporting evidence presented for efficacy should warrant not only the risks of the intervention but also other burdens on research participants imposed by the trial in general (29,30).

Nevertheless, some interviewees worried that increased attention to evidence for preclinical efficacy would delay clinical research, especially in cases where no in vivo efficacy data are available or useful due to a lack of model construct or predictive validity. There is no a priori reason for why this aspect should clash with strengthening the robustness of preclinical evidence for efficacy; if there is no point to conducting animal studies, they should not be conducted. Solutions for cases with no meaningful animal models could include pilot studies on a few patients, or sentinel dosing could limit the population subjected to high risk (31).

Greater attention to completeness was also a somewhat controversial topic. In general, attention to completeness was appreciated, but the interviewees were divided on which measures to apply and whether they were worth applying (21). As regulatory gatekeepers do not have the time and resources to check whether all studies that are relevant according to their definition are included, this gives sponsors much discretion in how they portray the supporting evidence. Of course, if this check became an explicit responsibility of regulators, they would then need to be supported in this.

To support regulators and RECs, some interviewees saw merit in presenting a clearer search strategy and transparent flow of included and excluded studies (32), for example, using visualizations such as bubble plots or flow charts (33). This was especially appreciated against the backdrop of concerns about publication bias in preclinical research (34–36). That said, some metaresearchers and ethicists whom we interviewed noted that preregistration would be the most effective measure for assuring the completeness of preclinical evidence. Regulatory guidance on IBs would need to require the preregistration of all included preclinical studies intended to directly inform decisions on launching clinical trials. This requirement would be relatively easy to implement, and non-preregistered evidence in IBs could be explicitly flagged. Additionally, existing registries for animal studies already allow embargo times, protecting intellectual property.

Finally, changes to the IB or the process of reviewing it can either increase or decrease administrative burden. It is therefore important to implement changes carefully and involve relevant stakeholders early. Our findings point to some potential blind spots in regulatory oversight, and further discussion should determine how any of the measures explored could be implemented in a way that would make them be beneficial to the stakeholders involved.

## Limitations

One limitation of our study is the selection of some interviewees by initial recommendations and subsequent “snowballing,” which could lead to an overrepresentation of participants who are more familiar with issues of reproducibility in preclinical research. Additionally, regional specificities in regulatory frameworks and the self-perception of RECs in different regions require a trade-off between a multiregional limitation of this study’s message on the one hand generalizability on the other. Last, our study is likely biased toward the European regulatory landscape. Although we made efforts to reach non-European regulators through multiple channels, none were available to participate.

## CONCLUSION

Currently, the responsibility to assure that the supporting evidence is complete and sufficiently robust lies with the sponsor, with few checks by regulatory and ethics gatekeepers. Possible measures to address complete reporting of the available evidence include the transparent reporting of the evidence synthesis strategy in a systematic review-like fashion or the preregistration of preclinical studies. Possible measures to address the issue of a lack of information on robustness include the reporting of key internal validity items for key studies in tabulated form (11). Our interviews revealed stakeholder reservations about these measures; some questioned whether they would be useful, and many were worried about overburdening the review system. In principle, having more robust decision-making processes in place aligns with the interests of all stakeholders and with many current initiatives to increase the translatability of preclinical research and limit the conduct of uninformative or ill-justified trials early in the development process. Further research should investigate which measures could be implemented to benefit all stakeholders.

## Data Availability

Raw data (interview transcripts) cannot be shared as participants were ensured that only members of the study team had access to transcripts as a condition of participation. A breakdown of the interviewees′ roles and sample quotations are available as supplementary materials.

## FINANCING AND CONFLICTS OF INTEREST

This study is financed by BIH QUEST departmental resources. The authors declare no conflicts of interest.

## ACKNOWLEDGMENTS

We thank Dr. Ulf Tölch, Dr. Natascha Drude, Dr. Alexandra Bannach-Brown, and Dr. René Bernard for their help with refining the interview guide by participating in pilot interviews and Johannes Schwietering and Dr. Sören Sievers for their feedback on earlier presentations. We thank Dr. Natascha Drude, Dr. Vince Madai, Dr. Benjamin G. Carlisle, Martin Holst, and Alice Faust for their comments on the manuscript.

## DATA AVAILABILITY

Raw data (interview transcripts) cannot be shared as participants were ensured that only members of the study team had access to transcripts as a condition of participation. A detailed breakdown of the interviewees’ roles and sample quotations are available as supplementary materials.

## AUTHOR CONTRIBUTIONS

Martin Haslberger: conceptualization, investigation, data curation, writing – original draft preparation, project administration; Susanne G. Schorr: conceptualization, writing – review and editing; Daniel Strech: supervision, conceptualization, writing – review and editing; Tamarinde Haven: conceptualization, methodology, investigation, writing – original draft preparation.

## Supporting Information

Table S1: Interviewees per stakeholder group and country (explanations in text)

Table S2: Selected quotations for SWOT – completeness

Table S3: Selected quotations for SWOT – robustness

