## Supplementary Materials for "Preclinical Efficacy in Investigator’s Brochures: Stakeholders’ views on measures to improve Completeness and Robustness"

Table S1: Interviewees per stakeholder group and country (explanations in text)

| **Region** | **Regulator** | **REC member** | **Ethicist/Meta** | **Industry** | **Total** |
|---|---|---|---|---|---|
| Germany | 3 | 2 | 1 | 3 | 9 |
| Netherlands | 1 | 4 | | | 5 |
| UK | 1 | | 2 | | 3 |
| USA | | 1 | 2 | | 3 |
| EU | 2 | | | | 2 |
| Belgium | 1 | | | | 1 |
| Canada | | | 1 | | 1 |
| Denmark | 1 | | | | 1 |
| France | | | | 1 | 1 |
| Switzerland | 1 | | | | 1 |
| **Total** | **10** | **7** | **6** | **4** | **27** |

Table S2: Selected quotations for SWOT – completeness

| # | SWOT | Citation |
|---|---|---|
| 1 | Justification of evidence selection | "Making it transparent and at least in principle reproducible so that somebody can say, OK, well, what did you search? How did you, what did you exclude? What gets included? How did you break this down? So I can be confident if I'm on the receiving end of this, that this is a valid story or this is appropriately grounded. And even if I may disagree with the conclusion, right, I nevertheless have enough information to understand, OK, well, here's how they got to where they are." – US medical ethicist |
| 2 | Justification of evidence selection | "Having been in industry myself, sometimes you wonder why a certain compound is chosen over another one or why a program in its entirety decides to go ahead versus another one. And the choices of that may be due to everything else but scientific consideration. So a program may be pushed, even if he's a bit shaky for the grounds, for other reasons. And this interplay between hard science and commercial consideration is, of course, unfortunately, yeah, it does take place and informs a lot of the decisions, even in what to show to the investigator." – EMA division head |
| 3 | Justification of evidence selection | "It's more about justifications, explanations. Sometimes the information you get might be just five sentences too short that you can actually accept the kind of argument. But I would say, with regard to in vivo studies, we hardly ever ask for in vivo studies in that scenario. There might be some in vitro cell-based assays we would ask for, but in general, it's the justifications." – Nonclinical assessor at a European national regulatory agency |
| 4 | Mitigating selective reporting | "If the preclinical evidence is insufficient, biased, stressing only the positive studies, leaving the negative studies aside, then that is a capital sin, I would say. Really putting subjects, human subjects, into grave danger so that that is something that the pharmacologist in the IRB together with one or more colleagues should be really, really … how do you say that we should really assess that very thoroughly." – US medical ethicist |
| 5 | Mitigating selective reporting | "I think it's a legal requirement that they have to present also not-so-good data. Can we verify that when they don't publish it? No. We can't, so I think it's stuck with the sort of moral, legal requirement that you would have to present this data." – Nonclinical assessor at a European national regulatory agency |
| 6 | Mitigating selective reporting | "It's a black box. You know, the criteria for preclinical development, toxicology studies, animal studies. And they are [unclear] by definition and by the EMA regulations. You have to do it; it's mandatory. You have a program. And what you cannot judge if there's any study, like, is there any study that has been done missing? It's impossible to prove for me, to check, you know, if they have a ... I cannot check this. So you have to trust that that's complete." – Clinical pharmacologist, IRB head |
| 7 | Mitigating selective reporting | "I had the impression that the information is always complete. So I was not aware of a problem there. And most of the studies originate from the pharmaceutical industry. So the question is, how can I be aware that it's not complete? Because most data belong to the pharmaceutical industry. And I'm only aware of what they report to me." – Hospital pharmacist, IRB member |
| 8 | Justifications are more important than complete data | "It's always a problem, that's when we get that kind of question like, what can we improve, in different meetings, for instance, I have attended, like, especially also for the IB. What is the most important part? It's a case-by-case decision. It's really always a pity to say this, but it's true. So what I said is there is no one-size-fits-all guideline, or maybe it always depends on your data and how you present the data and explain." – Nonclinical assessor at a European national regulatory agency |
| 9 | Justifications are more important than complete data | "I think you have to differentiate between two things. The one is the safety studies where you have clear guidelines, what should be available. So I think in that scenario, the completeness is more or less easy to be fulfilled. On the other hand, with regard to the efficacy studies, where you don't have clear guidance, but a lot of publications maybe or university studies, academia studies, there, in terms of completeness, it really depends. It's a case-by-case decision. Do you have one mode of action? Is there only one proof of concept? Is there a good animal model that could actually be used for it? So with regard to that, that's certainly more difficult." – Nonclinical assessor at a European national regulatory agency |
| 10 | Confidentiality | "The caveat you might face is, of course, that the amount of information that is available in the public domain might be limited. It depends a bit on your target and what you're looking at. But this is, of course, the difficulty you face when it comes to drug discovery work that is highly innovative. You possibly don't see a lot of, uh, literature on the topic itself. So that's an issue." – Scientific director at a pharmaceutical company |
| 11 | Confidentiality | "It's a bit of a problem; we can't broadcast.... We send in strict confidentiality to the health authorities a full package which is assessed by them, and they say, yes, the evidence is sufficient for a clinical trial to go." – Research quality manager at a pharmaceutical company |
| 12 | Confidentiality | "But that's not available for, let's say, an IMP that goes into a first-in-human because there are no published data, usually, the minority. Most of them are in the domain of confidentiality of, let's say, the pharmaceutical industry. So they should systematically review and report all the evidence that's there, but it's not in the public domain?" – MD, clinical researcher with previous industry positions |

| | | |
|---|---|---|
| 13 | Traceability of studies | ”[…] Try and make it transparent, forcing them to register the studies that they intend to do in advance – those studies in an international register. Then they report on those studies and whether those studies were abandoned and which ones they did do. And they give an assessment of the data. And perhaps as somebody referees it, you know that there's some quality assurance in the process. But, you know, governments need to take a role here in ensuring accountability.” – MD and metaresearcher |
| 14 | Traceability of studies | “But it's always important that the ongoing studies are also implemented in the IB. And this is sometimes, or maybe I'm answering already some of the questions you may have. From my point of view, a problem that occurs or a deficit, sometimes, in the IB is that the ongoing studies are not fully implemented yet because of the development process of the studies. There are ongoing studies that the IB has to be, there's a deadline has been, basically, there's a last-version draft of an IB, then the process of submission to the ethical board, global studies around the world, or the different centers, you know, is issued. And then there are studies ongoing, but they are not instantaneously, you know, filled, all of the information is filled in the IB. Actually, that could be something that's, I think, a potential issue or matter of improvement, you know, with being everything, being online, available and simultaneously that kind of a platform where an IB can be also, you know, updated continuously.” – MD, biometrician, REC chair |
| 15 | Economic benefits | “A good review of that is such a positive proposition economically because it would stop invalid research from going forward, it would stop huge wastes of money on both institutional sides, in terms of the waste of exposure, of the ethical exposure of humans, but in terms of money for pharmaceutical companies and for academic institutions and for the National Institutes of Health, it would stop the funding of uninformative trials.” – US IRB chair and CEO |
| 16 | Economic benefits | "My role was to counsel them on prospects of going into clinical trials, venturing into, I mean, assessing the portfolio basically, and then making a decision on whether they should go into it or not, among many other, obviously many other experts that gave their advice. And also in this capacity, I was involved in investigator brochures because obviously, what they were collecting and putting together at some point, sooner or later, when they made a decision to go into these investigator brochures. And for the record, I should say that I've never recommended going into clinical trials with what they had. I once made a calculation that if they would have listened, they would have saved, like, two billion.” – MD, translational researcher |
| 17 | Overburdening review bodies | “Everybody's overworked; they don't get enough support for the REC work that they're doing. And now we're asking them to do even more and make their committee even more political than it already is because, you know, it's going to be harder to get through that door than it currently is. And people already think it takes too long and it is too hard.” – US medical ethicist |
| 18 | Overburdening review bodies | “When an applicant writes a huge IB and presents a lot of data and then sometimes I ask myself if it's on purpose, because if you like to, yeah, make the regulator and the assessor a target, then you have not the right focus anymore, like you are reading and reading and lose the right focus because you get tired by reading all the data, which are not really important to find the important spot. So that makes more sense that they condense this written text, for instance, and focus on the right targets.” – Nonclinical assessor at a European national regulatory agency |
| 19 | Slowing innovation | “But as I'm not familiar with assessing IBs with this information, it could also be like a show-stopper because I would be confused if I certainly opened up and got a holistic insight on all the studies.” – Advisor at a European national regulatory agency |

### Table S3: Selected quotations for SWOT – robustness

| # | SWOT | Citation |
|---|---|---|
| 1 | Importance of IV | “It's not at all clear that the evidence reviewers would need to have access to validate efficacy inferences, in fact, you know, that they're able to do that. And if you can't assess the potential for efficacy, how in the world can you look at any ratios between risks and benefits?” – US medical ethicist |
| 2 | Importance of IV | “What could be improved, I think, is maybe a standardized way of documentation in terms of descriptive statistics and how would you present the data, etc., and also replication of experiments.” – Clinical pharmacologist, IRB member |
| 3 | Importance of IV | “Very often, as you put down, the robustness of the proof of concept may not be that strong. That's correct. […] It doesn't play a very big role for us. As you may be aware, there are no guidelines in support of that I could claim for the company to have done that robust research. Actually, there are some in the newest guideline from the EMA, the clinical trial and first-in-human that points the companies to that we do want to see some efficacy studies. But otherwise, I have nothing to back me up in such a claim.” – Advisor at a European national regulatory agency |
| 4 | Importance of IV | “Efficacy data that's leveraged for clinical trial approvals or for marketing authorization approvals in the absence of clinical trials isn't subjected to any regulatory framework. So there is no GLP for "we gave dogs with Duchenne dystrophy this drug"; that doesn't |

| | | |
|---|---|---|
| | | exist. So I don't think that the GLP for safety is fit for the purpose because it doesn't deal with things like randomization, blinding and the like, and there isn't any GLP for efficacy.” – Translational researcher, advisor at a national regulatory agency |
| 5 | Increasing accountability | “And we do sometimes see studies included that did not show what they expected, but then often they just include more essays or animal models to underscore their hypothesis. But most often they bring forward what has shown effect, as they are looking for. So it is certainly biased in that way.” – Chief advisor at a European national regulatory agency |
| 6 | Increasing accountability | “It's a giveaway; you can see if they start searching for many models, also in the tox part, that there may be a pointer here that it's not working. So for me, it could be a very nice thing to see.” – Chief advisor at a European national regulatory agency |
| 7 | Current practices suffice | “In general, we know that from time to time, there are, of course, cases when drugs entering phase one, they were not safe. Yeah, that's true. We cannot fully derisk new chemical entities during nonclinical development. That's clear. But overall, I think the system ... I'm not ... I'm open to anything new, but overall, the system works pretty well.” – Pharmacologist and CSO of a biotech company |
| 8 | Current practices suffice | “We see a lot of scientific advice on first-in-human trials, and this guides the process and ensures robustness at the moment that they come for the clinical trial application. Inversely, when we see a really lousy application, we can stop it and say, come back to us, but come first for scientific advice, and then you come back with your clinical trial application. So we have a dialogue with the sponsors if we have issues regarding robustness of information. But it's not ideal.” – Nonclinical assessor at a European national regulatory agency |
| 9 | Current practices suffice | “We could request that. But to be realistic, we would only do that if we had a great concern because the time and resources are not set for us to look at the original study reports. […] That actually makes it very important that the applicant draws out the most relevant assays and points in the whole IB. And we do rely on that. So I think after going through a lot of investigator's brochures, you kind of get the feel, when should I dig into something? When is this proof of concept as we speak of today a little weak or not really proven? Did they forget something?” – Chief advisor at a European national regulatory agency |
| 10 | Current practices suffice | “The fact is these clinical trial assessments are very short timeline assessments. We are having deadlines […] for phase one and first-in-human clinical trials of 15 calendar days. In that time frame, it's very difficult to review full study reports, as you can imagine. It's not possible. So what do we do? We review the summarized information in the IB. If we have a concern, we formulate a ground for nonacceptance or a request for further information in a pilot, according to the clinical trial regulations. And then the applicant can give us extra information on the specific topic. We will seldom request full study reports because, well, the time for review is just too short.” – Nonclinical assessor at a European national regulatory agency |
| 11 | Trust in sponsors’ incentives | “In general, I think that the pharmaceutical industry is trying to be very critical about the drug and therefore also be very precise about reporting the preclinical information because nobody wants to advance a drug that has no future. […] I think that that in itself creates a basis of trust.” – Clinical pharmacologist, REC chair |
| 12 | Utility of preclinical efficacy studies | “That's a little bit, you know, the problem that, yes, you disclose, and then you get very little input because the different viewpoint and even depth of knowledge that a clinical investigator may have, although these, you know, models that are nowadays enormously complicated, you may have access but say nothing of value. So it's always a game of trust.” – EMA nonclinical assessor |
| 13 | Utility of preclinical efficacy studies | “I've seen many times when I was in preclinical research and as well as in the regulator's shoes that there is a blissful ignorance of the investigator of what do they make out of this data, preclinical data and toxicology data. So they see all these EC50 and IC50s and, you know, strange models with the specificity, sensitivities. And for them, that doesn't mean anything, so they rely on the conclusion of the sponsor. And the sponsor has a lot of leeway in making the conclusion that they want. And they rarely are questioned.” – EMA division head |
| 14 | Utility of preclinical efficacy studies | “Preclinical evidence, very much like human pharmacology, is rarely bothered by statistical concerns. And that is because what you're looking for are the predictable things about the drug. […] It's much more about understanding than about confirmation when you don't know. So I'm not bothered by whether there's enough animals. I mean, if they all show the same response, then it doesn't matter if it's 6 or 10. If you do six, and two show the response, or three, then there's something wrong.” – Clinical pharmacologist, REC chair |
| 15 | More thorough assessment | “I don't want to make parallels to GLP, but if you say that a study was done, according to OECD 406, then you know exactly what it means. […] Also, for the agency, when they review them, they would know immediately which studies have been conducted with a greater rigor and which studies were not.” – Pharmacologist and CSO of a biotech company |
| 16 | More thorough assessment | “I mean, a radical thing would be to, say, only have a quality bar, and only a certain level of quality can even get into these things, but then you probably wouldn't; they would have a, you'd have to do a lot more experiments before any new decisions are |

| | | |
|---|---|---|
| | | made. I don't think it's that bad of an idea, to be honest." – Academic preclinical/metaresearcher |
| 17 | More thorough assessment | "If you're on an IRB, like you would ideally expect to get, right. With the protocol, here's the prior evidence, here's the risk of bias or quality assessment of that evidence. Here's what they found. Right. And again, it's like if you can just make it accessible for people to drill down a little bit where they have questions, I think that would be huge, hugely beneficial." – US medical ethicist |
| 18 | Interoperability | "Having the ability to sort of say, like, OK, well, show me the quality scores for these studies. Right. And that's kind of going down a level. Right. You can kind of see. OK, so the most relevant studies were mixed quality, but there was one that was really good. Right. And that's the one that's kind of really driving this. Well, let me click through there and actually call up the paper itself, right, and now I can read about this, right?" – US medical ethicist |
| 19 | Interoperability | "We are in the 21st century, so maybe IBs should have not only old-style references to reports, but those should be hypertext links. And then you just go straight to some secure online cloud storage, and then you would just be able to access and read the full report." – US medical ethicist |
| 20 | Interoperability | "So I think the key to that, to better and more complete data, is actually making it more simple to have all the data in one spot, in the same place and with an interface that allows the professionals that work with it, be they on the industry side, on the regulatory side, and, of course, all the people involved in ethical committees to have a way more interactive way of working with data." – EMA scientific advice officer |
| 21 | Interoperability | "The only thing where I think it would be easiest for regulators is if all the documentation that is mentioned, listed, is all also available for the regulator. So that I don't have to look up this paper because it's part of the submission, more or less. I think that would be very easy because obviously, the sponsor has all the information, so why not just compile it and put it on a CD-ROM or whatever, USB stick, some portal, however your agency works, and make it available? I think that it's then up to the agency to decide how deep you can go into it based on your timelines, your resources and the time you can invest. But at least it would be available." – Deputy division head at a European national regulatory agency |
| 22 | Protection of research participants | "There's a real divide when human researchers look at, or doctors look at, animal data; I think their brains look at it differently than they would for actual clinical studies. It's a different calibration, and I think they're much less likely to be skeptical of the data that's presented." – US IRB chairman and CEO |
| 23 | Protection of research participants | "We do have discussions, especially with new members in the team, that the most critical parts where you maybe have the volunteers is compared to marketing authorization, where you have patients and you have a lot of clinical trials data, and it's really this fast two-week business just based on this IB. Sometimes you have study reports depending on the phase; sometimes you don't. This is really, it feels like the wrong way. You should invest more time there and have more information to be sure the clinical trial participants are protected, compared to a marketing authorization, where you have so many clinical trials data that the nonclinical part is very limited, so the impact is limited." – Deputy division head at a European national regulatory agency |
| 24 | Protection of research participants | "I think there needs to be sort of informational justice or parity, for lack of a better term, between research participants on the one hand and investigators on the other, such that research participants who were so inclined can have access to information that will give them a robust understanding, right, of just what the predictive value of preclinical data are, right?" – US medical ethicist |
| 25 | Overburdening review bodies | "The institutional review board doesn't have the time or resources to double-check the scientific materials that are provided as part of the background. We take those for granted and sort of assuming those, we feel our job is to assess whether the research as presented to us is ethical, but that comes out of the history. I mean, the reasons the research regulations in the United States were passed were not because of lack of scientific rigor. They were passed because of complete blind spots about human subjects protection. So that's what IRBs are equipped to deal with." – US IRB chairman and CEO |
| 26 | Overburdening sponsors | "I don't think that we will ever have really strict guidelines on efficacy studies because it's research based, and this will hamper research if we say you have to conduct it always in a standardized way." – Nonclinical assessor at European national regulatory agency |
| 27 | Overburdening sponsors | "I do understand the ethical concern; I do understand. But, you know, I don't see. So what's the duration that would make ... I mean, is it "do more studies up front and come later for first-in-human trials," that would be the only proposal. And I can assure you, you will have discussions, I would guess, with animal protection activists for sure, because even if it's not working, then you wasted all these animals." – Deputy department head at a European national regulatory agency |
